# Multidimensional Sleep Health: Concepts, Advances, and Implications for Research and Intervention

**DOI:** 10.1101/2021.04.20.21255799

**Authors:** Joon Chung, Matthew Goodman, Tianyi Huang, Suzanne Bertisch, Susan Redline

## Abstract

The new paradigm of multidimensional sleep health (‘sleep health’) offers both challenges and opportunities for sleep science. Buysse (2014) has described sleep health to be multidimensional, framed as positive attributes, operationalizable into composite measures of global sleep health, sensitive to upstream exposures, and consequential for downstream health. We highlight two paradigm-shifting effects of a multidimensional sleep health perspective. The first is the use of composite sleep metrics which i) enable quantification of population shifts in sleep health, ii) with possibly reduced measurement error, iii) greater statistical stability, and iv) reduced multiple-testing burdens. The second is that sleep dimensions do not occur in isolation, that is, they are commonly biologically or statistically dependent. These dependencies complicate hypothesis tests yet can be leveraged to inform scale construction, model interpretation, and inform targeted interventions. To illustrate these points, we i) extended Buysse’s Ru SATED model; ii) constructed a conceptual model of sleep health; and iii) showed exemplar analyses from the Multi-Ethnic Study of Atherosclerosis (n=735). Our findings support that sleep health is a distinctively useful paradigm to facilitate interpretation of a multitude of sleep dimensions. Nonetheless, the field of sleep health is still undergoing rapid development and is currently limited by: i) a lack of evidence-based cut-offs for defining optimal sleep health; ii) longitudinal data to define utility for predicting health outcomes; and iii) methodological research to inform how to best combine multiple dimensions for robust and reproducible composites.

## Introduction

Although it would be convenient if we had a single variable in our data sets called ‘sleep’ that encompasses the complex neurophysiologic process of sleep, what data sets typically include is a wide range of metrics *about* sleep: estimates of specific features of sleep as well as the impact of sleep on daytime symptoms and function. Each feature or *dimension* of sleep is often modeled separately and interpreted independently – as exposure or outcome – in distinct models: an effect size for total sleep time, one for sleepiness, and yet another for sleep efficiency.

However, sleep dimensions do not exist in isolation, and sleep health is multidimensional (1). Just as food is not consumed as singular nutrients (2), sleep is not experienced as singular dimensions. Adults who meet national recommendations of 7-9 hours of duration may evince vast heterogeneity in sleep latency, continuity, macro- and micro-architecture, and satisfaction. Moreover, a substantial body of evidence suggests that a variety of sleep dimensions influence academic outcomes, mental and physical health, illness, and mortality; and that these dimensions are sensitive to exposures at the individual level (*e*.*g. age; physiological shifts over the life-course, such as menopause; financial strain; caffeine, tobacco, and other substance use; physical activity, genetic risk loci, etc*.) and social and environmental levels (*e*.*g. neighborhoods and the built environment; social cohesion; social relationships; socioeconomic status in childhood; marital status; loneliness; discrimination; and work schedules, etc*.). Accordingly, Buysse (2014) has defined sleep health as: “a multidimensional pattern of sleep-wakefulness, adapted to individual, social, and environmental demands, that promotes physical and mental well-being” (1).

A multidimensional paradigm is, however, already intrinsic to our understanding of sleep. Clinicians, for instance, routinely consider multiple aspects of sleep in diagnosis and management of sleep disorders. Moreover, the literature has long reflected interest in the combined, including interactive, aspects of sleep, although has focused more on sleep disturbances rather than sleep health, and focused on clinical disorders rather than population health. Thus: What is distinctive and useful about the paradigm of ‘multidimensional sleep health’? How does it differ from prior and current work? Does ‘multidimensional sleep health’ constitute a paradigm shift, that is, a new *way* of thinking about sleep?

We provide empirical data to support multidimensional sleep health as a distinctively useful approach for characterizing ‘sleep health’ across the population. One innovation is the quantification of sleep ***health***, which ranges beyond disorder or insufficiency – acknowledging the existence of gradients of “healthy sleep” beyond meeting a minimum duration of sleep or an absence of disorders such as insomnia or obstructive sleep apnea (OSA) (1). Sleep health also parallels broader paradigm shifts which acknowledge the World Health Organization’s definition of health as more than the absence of illness (3).

While these are important frameshifts, sleep health has other distinctive but less emphasized features, which render it a more radical and useful paradigm shift than would seem at first glance. First, sleep health approaches multiple sleep dimensions (conceptually, operationally, analytically) as contributors to a unitary or a *composite* concept – a metric of global sleep health (1, 4). Extant scales include Buysse’s Ru SATED scale and the National Sleep Foundation’s Sleep Health Index (5, 6). Consistent and valid scales are necessary for scalable, longitudinal data collection that enables quantification of population shifts in global sleep health. Such work does not preclude analyzing each dimension in its own right, although statistical dependence among dimensions complicates model interpretation because sleep dimensions do not exist in isolation.

Second, sleep health emphasizes that sleep dimensions do not exist in isolation (4, 7). There are physiological reasons why certain dimensions are correlated, perhaps causally, with others, for instance, duration and sleepiness, OSA and continuity, timing irregularity and duration, circadian body-temperature nadir and sleep onset latency, et al. (8-11). Sleep dimensions also may show statistical correlation when derived from common measurement tools. Thus, interpretation of individual dimensions as if they were independent may lead to erroneous, or partial conclusions about ‘sleep’: what kind of continuity, quality, alertness, and regularity is likely to be seen among ‘sufficient’ sleepers (however defined) as opposed to ‘insufficient’ sleepers?

The purpose of this study was to illustrate these two frameshifts: i) unidimensional composites; and ii) consideration of correlations. We explored the utility of expanding Buysse’s model by incorporating sleep stage information (% N3, % Rapid Eye Movement [NR]), the Apnea-Hypopnea Index[AHI], self-reported frequency of long sleep onset latency (an insomnia complaint), and regularity in duration (12, 13) by mapping these dimensions to a conceptual sleep health model. We considered how to model multiple dimensions and found merit in both analyzing individual dimensions as well as a global sleep health metric: from a public health perspective, it was important to know which sleep dimensions are the drivers of a global sleep effect. Finally, we showed how these composite and individual dimensions map across race/ethnic groups, an extension of prior work (14).

## Methods

### Sample

The sample was from the Multi-Ethnic Study of Atherosclerosis (MESA), a 6-community cohort of aging adults, diverse in race/ethnicity (White, Black, Hispanic, Chinese). Details on this cohort are published (15). In brief, at Exam 5 (2010-2013), participants were invited to the MESA-Sleep ancillary study and underwent single-night at-home polysomnography (PSG), 7-day wrist actigraphy (Actiwatch Spectrum; Philips Respironics, PA; Actiware-Sleep v 5.59), and validated sleep questionnaires (Epworth Sleepiness Scale, Women’s Health Initiative Insomnia Rating Scale) (16, 17). A previous racial-ethnic sleep disparities project included adults 54-93 years old (14); in following analyses, only adults 54-64 years were included, because there are more normative data available for this age range (18).

### Sleep health conceptual model

As described previously (14), sleep health domains were drawn primarily from Ru SATED but also the National Sleep Foundation’s Sleep Health Index (6). Selection of additional variables was informed by prior knowledge of a high prevalence of OSA and sleep fragmentation in middle-age and older adults (19). Thus, several continuity metrics were chosen *a priori*: sleep maintenance efficiency (SME), the Fragmentation Index (FI), and Wake after Sleep Onset (WASO). Other additions to Ru SATED such as % NR, % N3, AHI (3% desaturation to define hypopneas), duration regularity, and self-reported frequency of un/desirable onset latency (freq. of difficulties initiating sleep) were also selected *a priori*, using expert knowledge or evidence from the literature indicating their relevance for health outcomes (20). Specifically, the mapping of each sleep variable to our conceptual model of sleep focused on these four periods (Figure 1): 1) the transition from wake to sleep (onset latency, self-report) and daytime sequalae (quality, alertness) from the participants’ perspective; 2) the period between sleep onset and offset evaluated objectively by PSG (eg, sleep architecture and/or actigraphy (eg, fragmentation); 3) inter-daily variability in timing and duration; and 4) the entire sleep experience both during and across nights (14).

**Figure 1.**
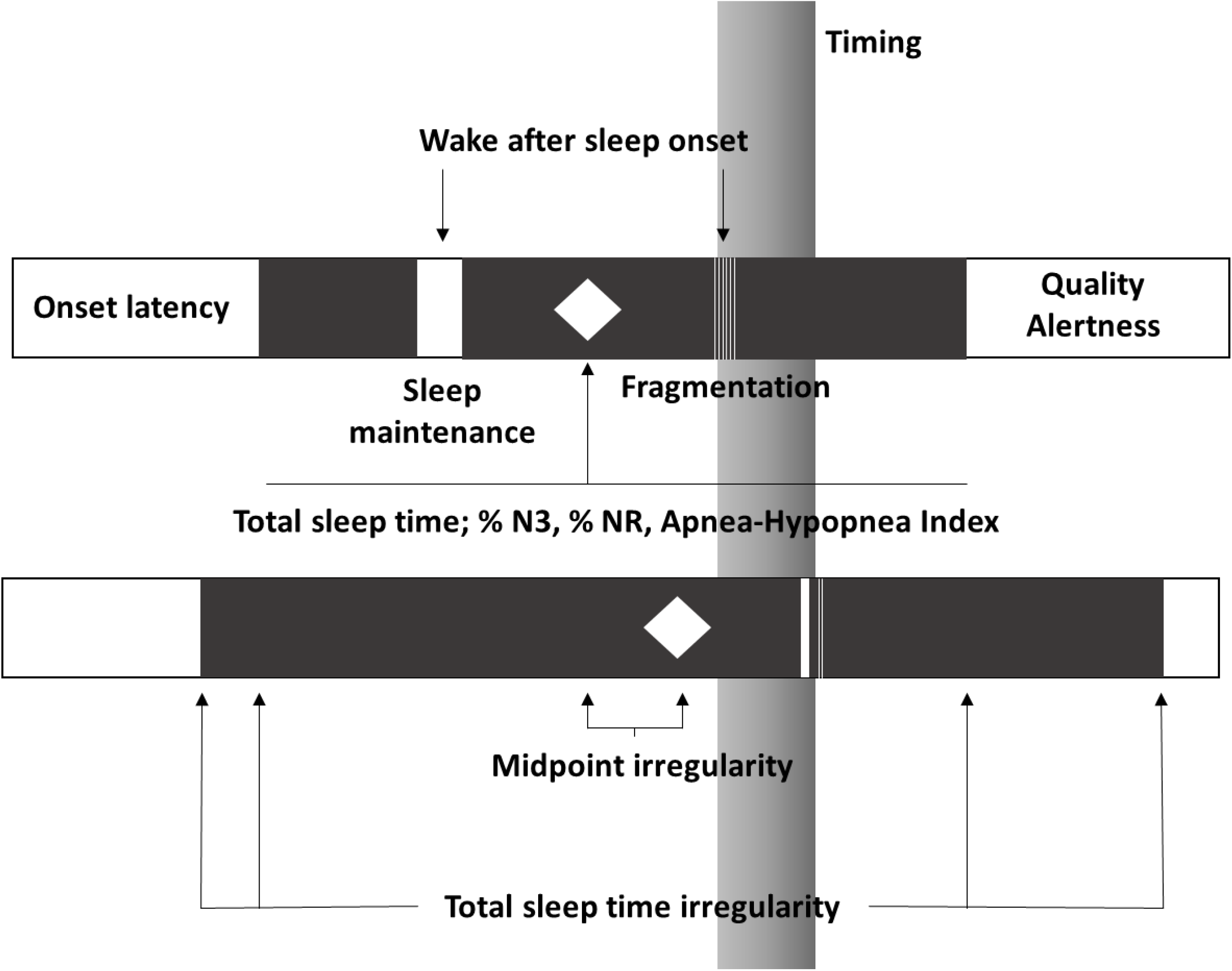
Conceptual Sleep Health dimensions across time. Note: Diagram of two consecutive days/nights of sleep. Dark bars indicate sleep; white bars indicate wake. This diagram is conceptual and does not represent a particular individual in MESA-Sleep. Diamonds indicate midpoint. Onset latency, quality, and alertness are evaluated during wake (self-report).

Two strategies were used to construct sleep health composites: i) Principal Components Analysis (PCA); and ii) a summary score of dichotomized sleep variables. PCA was conducted on a) Ru SATED variables (midpoint regularity [MPsd], quality, timing (log-transformed), maintenance efficiency, and duration); b) Ru SATED, expanded with selected variables from PSG, adding AHI, % N3, %NR; c) all variables chosen *a priori* for the Sleep Health Score (adding several variables measuring similar dimensions: duration regularity, sleep onset latency (frequency of trouble sleeping), WASO, fragmentation) (14). The resulting PC scores were standardized and coded so that higher values represent better health, and a 1-unit increase corresponds to a 1-sd increase in sleep health. For PCA, actigraphy-assessed onset latency was used rather than self-report due to the high skewness of the latter ordinal variable.

As described in (14), for the summary score (21-23), cut-points defined optimal ranges, with coding of ‘favorable’ or ‘healthier’ sleep as ‘1’ and non-optimal ranges as ‘0’. Optimal ranges were drawn from the literature, the NSF’s objective sleep quality report (eg, for WASO), expert consensus, or sample characteristics (12, 13, 18, 19, 21). These dichotomous indicators of favorable sleep were summed into a global metric, a Sleep Health Score (SHS), additionally categorized as: least favorable (SHS 0-3), less (4-6), more (7-9), and most favorable (9-12) sleep health.

### Statistical Methods

Pearson correlations were computed for continuous sleep metrics (log-transformed where necessary). Correlations were further investigated by inspecting Principal Components for three composites: (1) Ru SATED; Ru SATED + OSA/Sleep Architecture; and the Sleep Health Score (comprehensive indices across domains). Sensitivity analyses assessed a parsimonious version of the Sleep Health Score (PC1) that eliminated potentially redundant measures. Composite score internal reliability was assessed by alpha Cronbach. Consistency in PC weights - both direction and magnitude - was evaluated across each component. Trends among individual sleep health variables (e.g., duration, sleepiness) in relation to SHS categories were reported. Global sleep health variations by racial/ethnic group were assessed for potential utility for sleep health disparities research in the non-elderly. Outcomes of composite sleep health scores (linear regression), and their dichotomized components (modified Poisson regression (24)), were regressed on the exposure of race-ethnicity (White=ref), with adjustment for age and sex. Analyses were conducted in R 3.6.3.

## Results

The sample of 735 ethnically/racially diverse participants had an average age of 59.4 ±3.0 years, 55.6% were female, 31.8% were currently employed, and most obtained at least a high school degree (n=664, 90.3%) (Table 2). The majority met criteria for actigraphy-assessed sufficient sleep (6-8 hrs; n=461, 62.7%; Table 1) but did not meet favorability thresholds for: continuity (WASO (6.8% favorable), fragmentation (27.1%)), sleep architecture (% N3 [11.4%], % NR [34.1%]), regularity (midpoint (21.4%) and duration (36.9%)), AHI (48.2%), and quality (21.4%).

**Table 1.** Sleep variables, dimensions, and dichotomization rules. The Multi-Ethnic Study of Atherosclerosis, age < 65 years (n=735).

| Sleep variable | Sleep health dimension | Measure | Dichotomization rule (coded as '1') | Sample prevalence of favorability ('1'). n (%) | Abbreviation |
| --- | --- | --- | --- | --- | --- |
| Midpoint variability (sd, min) | Regularity | Actigraphy | <30 minutes (13) | 157 (21.4%) | MPsd |
| Duration variability (sd, min) | Regularity | Actigraphy | <60 minutes (13) | 271 (36.9%) | TSTsd |
| Quality (WHIIRS Likert subscale; higher is increasing complaints) | Satisfaction | Questionnaire | "Sound and restful" or "very sound or restful" | 157 (21.4%) | Quality |
| Sleepiness | Alertness | Questionnaire | ≤10 (17) | 199 (27.1%) | ESS |
| Timing (midpoint)<sup>b</sup> | Timing | Actigraphy | 02:00-04:00<sup>b</sup> | 466 (63.4%) | Timing |
| Sleep Maintenance Efficiency (%) | Efficiency | Actigraphy | >90% | 524 (71.3%) | SME |
| Fragmentation Index<sup>a</sup> | Efficiency | Actigraphy | ≤15<sup>a</sup> | 199 (27.1%) | FI |
| Wake after Sleep Onset (min) | Efficiency | Actigraphy | ≤20 minutes (18) | 50 (6.8%) | WASO |
| Total Sleep Time<sup>b</sup> (min) | Duration | Actigraphy | 6-8 hours<sup>b</sup> | 461 (62.7%) | TST |
| %NR<sup>b</sup> | Architecture | PSG | 21%-30%<sup>b</sup> (18) | 251 (34.1%) | % NR |
| %N3<sup>b</sup> | Architecture | PSG | 16%-20%<sup>b</sup> (50) | 84 (11.4%) | % N3 |
| Apnea-Hypopnea Index (events/hr) | SDB | PSG | ≤15 events/hr (19) | 354 (48.2%) | AHI |
| Sleep onset latency (difficulties initiating sleep – WHIIRS subscale) | Wake-sleep transition | Questionnaire | "Less than once a week" or "no, not in the past 4 weeks" | 476 (64.8%) | SOLsr |
| Sleep onset latency (min) | Wake-sleep transition | Actigraphy | - |  | SOL |

a: The cut-point for FI was defined as the nearest whole number to the most favorable quartile (lowest quartile).
b: These dimensions have optimal ranges in which overexpression of this dimension is considered problematic.

**Table 2.** MESA-Sleep Participant socio-demographics, global sleep health, and sleep health metrics by Sleep Health Score. The Multi-Ethnic Study of Atherosclerosis, ages <65 years (n=735).

|  |  | Sleep Health Score categorized: least to most favorable SHS |  |  |  |  |
| --- | --- | --- | --- | --- | --- | --- |
|  | Overall | least (0-3) | less (4-6) | more (7-9) | most (9-12) | p |
| N | 735 | 120 | 378 | 205 | 32 |  |
| **Socio-demographics** |  |  |  |  |  |  |
| Race-ethnicity (%) |  |  |  |  |  | <0.001 |
| White | 265 (36.1%) | 26 (21.7%) | 123 (32.5%) | 96 (46.8%) | 20 (62.5%) |  |
| Chinese | 93 (12.7%) | 11 (9.2%) | 49 (13.0%) | 30 (14.6%) | 3 (9.4%) |  |
| Black | 200 (27.2%) | 52 (43.3%) | 108 (28.6%) | 38 (18.5%) | 2 (6.2%) |  |
| Hispanic | 177 (24.1%) | 31 (25.8%) | 98 (25.9%) | 41 (20.0%) | 7 (21.9%) |  |
| Female (%) | 409 (55.6%) | 55 (45.8%) | 205 (54.2%) | 126 (61.5%) | 23 (71.9%) | 0.011 |
| Age | 59.4 (3.0) | 59.3 (3.0) | 59.6 (3.0) | 59.2 (2.9) | 59.4 (2.9) | 0.638 |
| Education |  |  |  |  |  | 0.082 |
| Less than high school | 71 (9.7%) | 11 (9.2%) | 43 (11.4%) | 14 (6.8%) | 3 (9.4%) |  |
| High school or some college | 352 (47.9%) | 57 (47.5%) | 191 (50.5%) | 95 (46.3%) | 9 (28.1%) |  |
| College degree | 174 (23.7%) | 34 (28.3%) | 79 (20.9%) | 52 (25.4%) | 9 (28.1%) |  |
| Graduate | 138 (18.8%) | 18 (15.0%) | 65 (17.2%) | 44 (21.5%) | 11 (34.4%) |  |
| Married | 480 (65.3%) | 65 (54.2%) | 248 (65.6%) | 141 (68.8%) | 26 (81.2%) | 0.010 |
| Employed | 234 (31.8%) | 42 (35.0%) | 124 (32.8%) | 59 (28.8%) | 9 (28.1%) | 0.615 |
| **Global sleep health** |  |  |  |  |  |  |
| Sleep Health Score<sup>†</sup> | 5.7 (2.1) | 2.6 (0.7) | 5.1 (0.8) | 7.8 (0.8) | 10.3 (0.5) | <0.001 |
| Sleep Health Score (PC1)<sup>†</sup> | 0 (1) | -1.1 (0.9) | -0.2 (0.8) | 0.8 (0.5) | 1.4 (0.4) | <0.001 |
| Ru SATED (PC1)<sup>†</sup> | 0 (1) | -1.0 (0.9) | -0.1 (0.9) | 0.6 (0.7) | 1.2 (0.5) | <0.001 |
| Ru SATED + OSA + Architecture (PC1)<sup>†</sup> | 0 (1) | -1.2 (0.9) | -0.1 (0.8) | 0.7 (0.6) | 1.4 (0.4) | <0.001 |
| **Sleep Health variables** |  |  |  |  |  |  |
| **Regularity** |  |  |  |  |  |  |
| Midpoint variability<sup>a</sup> (sd, min)<sup>↓</sup> | 47.2<br>[31.8, 70.6] | 67.8<br>[47.9, 95.2] | 53.0<br>[36.9, 76.1] | 35.3<br>[25.5, 50.8] | 23.2<br>[16.7, 26.7] | <0.001 |
| Duration variability<sup>a</sup> (sd, min)<sup>↓</sup> | 76.4 (36.8) | 90.1 (29.9) | 83.2 (38.0) | 61.6 (32.0) | 39.6 (12.8) | <0.001 |
| **Satisfaction** |  |  |  |  |  |  |
| Quality<sup>c</sup> (Likert subscale<sup>d</sup>;<br>higher=increasing complaints)<sup>↓</sup> | 2.8 (0.9) | 3.3 (0.8) | 2.8 (0.9) | 2.4 (0.8) | 2.3 (0.7) | <0.001 |
| **Alertness/sleepiness** |  |  |  |  |  |  |
| Epworth Sleepiness Scale<sup>c↓</sup> | 6.3 (4.2) | 9.0 (4.9) | 6.2 (4.1) | 5.4 (3.4) | 4.1 (2.3) | <0.001 |
| **Timing** |  |  |  |  |  |  |
| Timing<sup>a</sup> (midpoint minutes from<br>midnight)<sup>↓</sup> | 196.1<br>[155.1, 242.5] | 230.8<br>[151.2, 278.8] | 201.1<br>[157.9, 254.6] | 182.1<br>[151.2, 222.3] | 177.5<br>[151.2, 205.1] | 0.001 |
| **Efficiency** |  |  |  |  |  |  |
| Sleep Maintenance Efficiency<sup>a</sup> (%)<sup>↑</sup> | 91.4 (3.4) | 89.2 (4.3) | 91.1 (3.1) | 93.0 (2.2) | 93.2 (1.8) | <0.001 |
| Fragmentation Index<sup>a↓</sup> | 19.3 (6.8) | 23.3 (7.4) | 20.3 (6.7) | 15.7 (4.7) | 14.7 (4.0) | <0.001 |
| Wake after Sleep Onset<sup>a</sup> (min)<sup>↓</sup> | 76.8 (56.3) | 102.8 (63.2) | 81.9 (58.5) | 57.3 (40.0) | 45.5 (33.1) | <0.001 |
| **Duration** |  |  |  |  |  |  |
| Total Sleep Time<sup>a</sup> (hrs)<sup>↗</sup> | 6.4 (1.2) | 5.6 (1.4) | 6.4 (1.2) | 6.9 (0.9) | 7.2 (0.6) | <0.001 |
| **Sleep Architecture** |  |  |  |  |  |  |
| % NR<sup>b↗</sup> | 19.0 (6.6) | 15.5 (6.9) | 18.4 (6.5) | 21.2 (5.7) | 23.8 (4.0) | <0.001 |
| % N3<sup>b↗</sup> | 9.7<br>[3.2, 16.7] | 8.2<br>[3.2, 14.7] | 8.6<br>[1.9, 15.9] | 10.9<br>[4.9, 17.6] | 18.6<br>[11.9, 20.0] | <0.001 |
| **Obstructive Sleep Apnea** |  |  |  |  |  |  |
| Apnea-Hypopnea Index<sup>b</sup> (events/hr)<sup>↓</sup> | 22.0 (20.2) | 34.6 (23.7) | 23.7 (20.0) | 13.6 (13.7) | 8.3 (6.7) | <0.001 |
| **Wake-sleep transition – insomnia**<br>**complaint** |  |  |  |  |  |  |
| Onset latency<sup>c</sup> (freq. of difficulties<br>initiating sleep) |  |  |  |  |  | <0.001 |
| No, not in the past 4 weeks | 405 (55.1%) | 33 (27.5%) | 203 (53.7%) | 142 (69.3%) | 27 (84.4%) |  |
| Yes, less than once a week | 71 (9.7%) | 6 (5.0%) | 28 (7.4%) | 34 (16.6%) | 3 (9.4%) |  |
| Yes, 1-2 times a week | 132 (18.0%) | 37 (30.8%) | 74 (19.6%) | 21 (10.2%) | 0 (0.0%) |  |
| Yes, 3-4 times a week | 66 (9.0%) | 17 (14.2%) | 41 (10.8%) | 7 (3.4%) | 1 (3.1%) |  |
| Yes, 5+ times a week | 61 (8.3%) | 27 (22.5%) | 32 (8.5%) | 1 (0.5%) | 1 (3.1%) |  |
| **Wake-sleep transition (actigraphy)** |  |  |  |  |  |  |
| Onset latency<sup>a</sup> (actigraphy) | 4.2 (3.0) | 4.6 (3.8) | 4.1 (2.9) | 4.1 (2.7) | 3.6 (2.1) | 0.326 |

↑ = Higher is better; ↓ = lower is better; ↗ = generally higher is better, but overexpression can be problematic (eg long sleepers, excessive % REM)
a: actigraphy
b: polysomnography
c: self-report
d: Women's Health Initiative Insomnia Rating Scale, subscale
p-values for skewed variables by Kruskal-Wallis; otherwise ANOVA.

Figure 2a shows the Sleep Health Score, Sleep Health Score (PC1), Ru SATED + OSA/Sleep Architecture (PC1), Ru SATED (PC1), and their components by race-ethnicity, adjusting for age and sex. Consistent with a prior report (14), the sample averaged 5.7 ± 2.1 of 13 possible favorable dimensions on the SHS. Blacks averaged 1.34 fewer favorable domains than Whites in global sleep health (p<0.001). Hispanics averaged 0.74 fewer favorable domains (p<0.001); however, most of the dimension-level disparities in Hispanics overlap with the reference. Ru SATED (PC1), Ru SATED + OSA/Sleep Architecture (PC1) showed similar variation by race/ethnicity as the Sleep Health Score, and Sleep Health Score (PC1). Figure 2b suggests that the largest drivers of global racial-ethnic disparities were sleep regularity (timing and duration), continuity (fragmentation, maintenance efficiency), and total sleep time.

**Figure 2.**
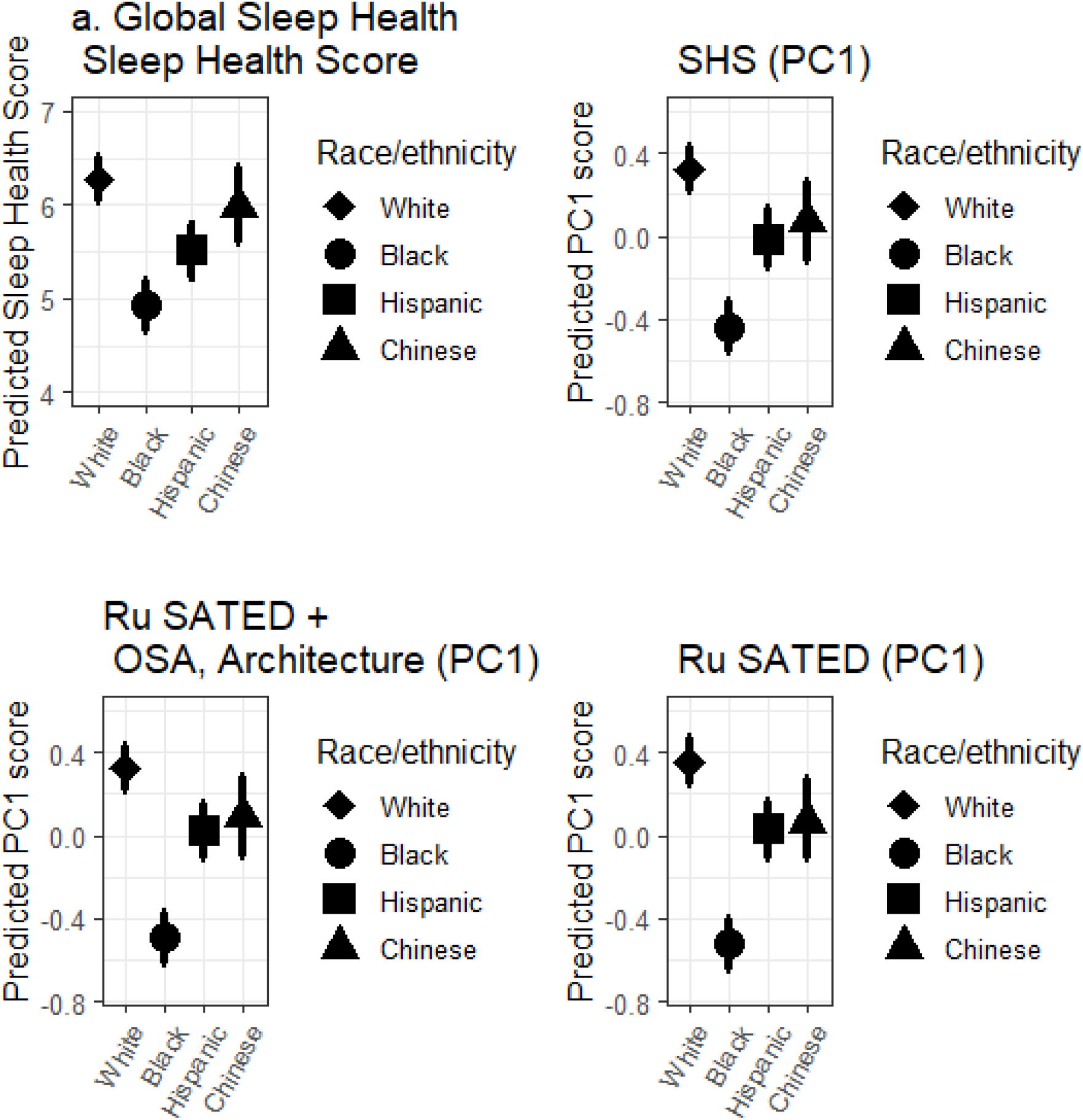
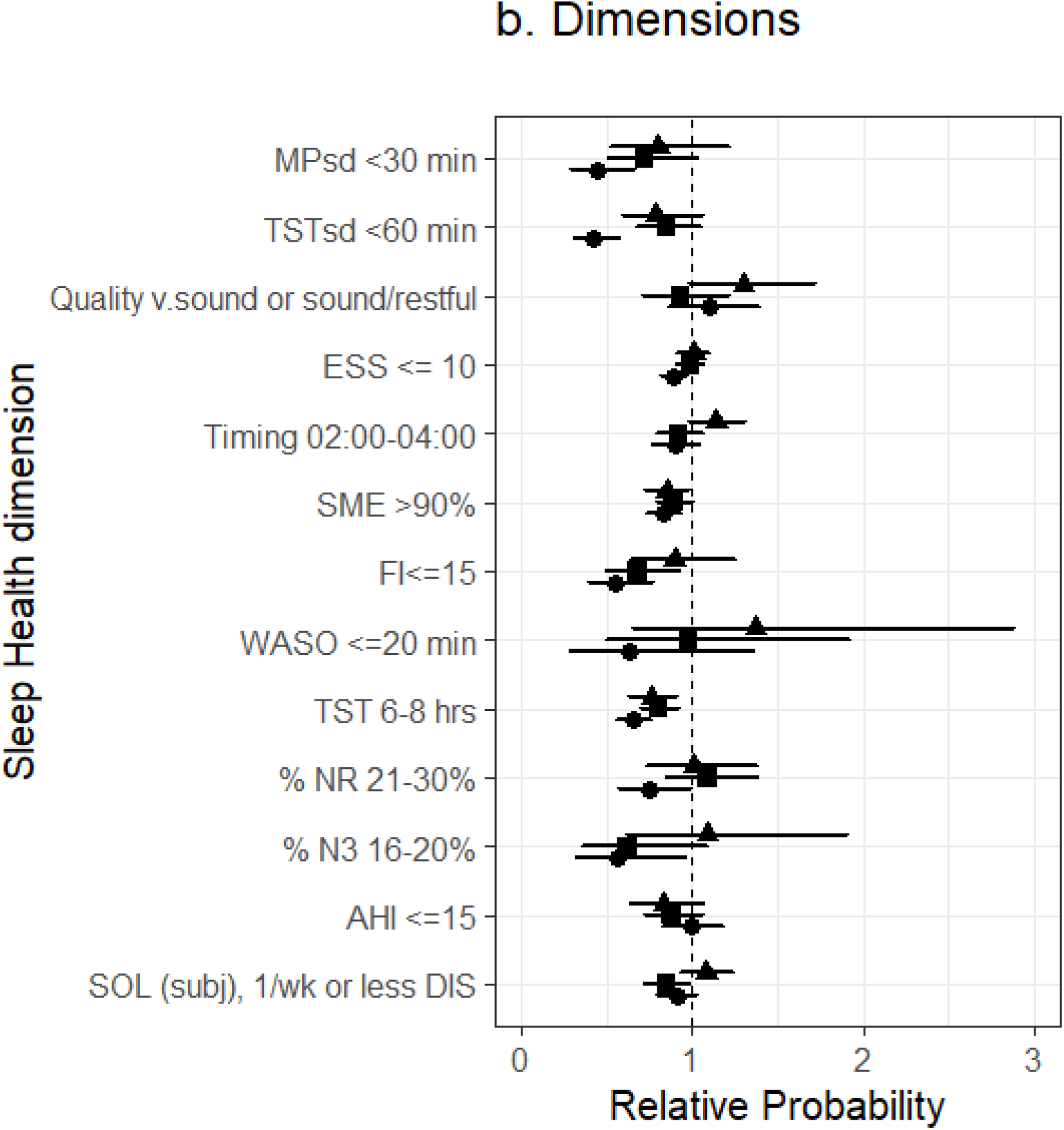
Age and sex-adjusted estimates of a) global sleep health and b) its components regressed on race-ethnicity. The Multi-Ethnic Study of Atherosclerosis (n=735).

Figure 3 shows correlations among sleep variables within and across domains. Within the continuity domain, fragmentation and sleep maintenance efficiency are highly correlated (ρ=0.72). Across domains, there are non-trivial statistical dependencies: timing regularity (MPsd(log)) and sleep duration (TST) are moderately correlated (ρ=0.40), as are timing regularity with sleep timing (Timing(log)) (ρ=0.32). AHI correlates with continuity metrics (FI, SME, WASO) and % NR (ρ’s range: −0.34 to 0.32).

**Figure 3.**
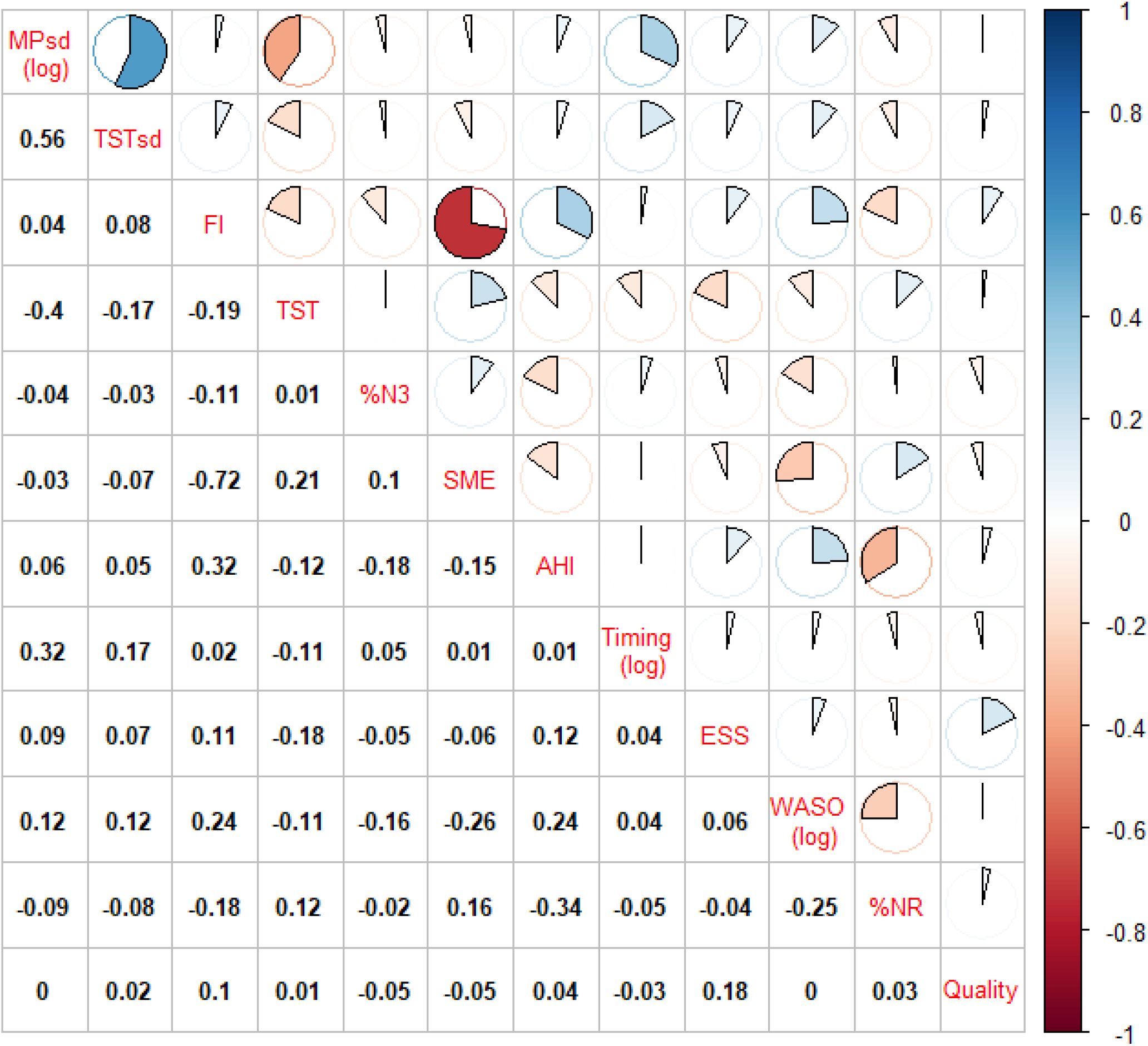
Pearson correlation matrix of objective sleep variables and Epworth scores. The Multi-Ethnic Study of Atherosclerosis (n=735)^a^. a. onset latency, self-report was omitted due to unimprovable skew. Quality is a Likert-scale, approximately normally distributed; lower is better. b. Pie size and shading indicate degree of correlation.

Table 2 suggests that when the components in Table 1 are summed into the SHS, increasing Sleep Health Scores capture variable yet systematic positive shifts across all sleep dimensions. Mean (median) differences in most-least favorable sleep health were notable in duration (+1.6 hours), midpoint regularity (−43.8 minutes), duration regularity (−48.7 minutes), AHI (−23.1 events/hr), % NR (+8.4%), % N3 (+9.9%), and WASO (−58.5 minutes).

Table 3 shows PC weights for the first three PCs for each composite score (variance explained ranged from 43.0% for the SHS (PC1) to 66.3% for Ru SATED). The first PC for each score is consistently interpretable in all models as a “sleep health score”: the directionality of PC1 weights for individual variables concur with *a priori* knowledge of “better” sleep (e.g., higher AHI contributes negatively, higher TST contributes positively). As expected, the PC1 weights varied depending on variables included but did not substantively change the interpretation of the PC1 composites. The simplest Ru SATED score shows that the measures of irregular timing, absolute timing, and duration loaded on PC1 while satisfaction/alertness and efficiency loaded on PC2. When measures from PSG and actigraphy were added to the Ru SATED score, AHI tended to load with efficiency and irregularity, %NR loaded with satisfaction/alertness, irregular duration loaded with irregular timing, and fragmentation loaded with efficiency. Across the extended (SHS) scores, timing and regularity tended to load on PC2 while self-reported sleep indices loaded on PC3. Moderate internal consistency was observed for different sleep health composites, with alpha Cronbach varying from 0.42 for Ru SATED to 0.61 for the full SHS.

**Table 3.** Principal Components Analysis of Sleep Health dimensions: Ru SATED, Ru SATED + OSA and Architecture, SHS, SHS-parsimonious.

|  | Ru SATED |  |  | Ru SATED + OSA and Sleep Architecture |  |  | SHS-PC1 |  |  | SHS-PC1 (parsimonious) |  |  |
| --- | --- | --- | --- | --- | --- | --- | --- | --- | --- | --- | --- | --- |
| | $\alpha=0.42$ | | | $\alpha=0.47$ | | | $\alpha=0.61$ | | | $\alpha=0.45$ | | |
| Proportion of Variance | 27.72% | 20.01% | 17.06% | 20.40% | 15.09% | 13.01% | 19.55% | 13.71% | 9.32% | 18.39% | 14.06% | 11.75% |
|  | PC1 | PC2 | PC3 | PC1 | PC2 | PC3 | PC1 | PC2 | PC3 | PC1 | PC2 | PC3 |
| **Ru SATED** |  |  |  |  |  |  |  |  |  |  |  |  |
| Irregularity<sup>↓</sup> (timing) | -0.59 | 0.26 | -0.12 | -0.45 | -0.45 | 0.06 | -0.31 | -0.53 | 0.11 | -0.45 | -0.43 | 0.10 |
| Satisfaction/quality<sup>↓</sup> (increasing insomnia symptoms) | -0.07 | -0.61 | -0.45 | -0.08 | 0.22 | 0.63 | -0.07 | 0.10 | 0.50 | -0.08 | 0.23 | 0.60 |
| Alertness<sup>↓</sup> (increasing ESS) | -0.31 | -0.51 | -0.31 | -0.29 | 0.10 | 0.53 | -0.18 | -0.03 | 0.37 | -0.29 | 0.10 | 0.50 |
| Timing<sup>↓</sup> (increasing delay) | -0.39 | 0.41 | -0.35 | -0.26 | -0.49 | -0.04 | -0.14 | -0.36 | 0.00 | -0.25 | -0.46 | 0.01 |
| Efficiency<sup>↑</sup> (maintenance) | 0.24 | 0.36 | -0.69 | 0.32 | -0.29 | 0.06 | 0.41 | -0.32 | -0.09 | 0.33 | -0.32 | 0.05 |
| Duration<sup>↗</sup> | 0.58 | 0.05 | -0.29 | 0.50 | 0.21 | -0.07 | 0.33 | 0.24 | -0.03 | 0.49 | 0.24 | -0.07 |
| **PSG (architecture)** |  |  |  |  |  |  |  |  |  |  |  |  |
| Apnea-Hypopnea Index<sup>↓</sup> |  |  |  | -0.38 | 0.43 | -0.23 | -0.32 | 0.18 | -0.29 | -0.38 | 0.38 | -0.27 |
| % N3<sup>↗</sup> |  |  |  | 0.14 | -0.34 | -0.18 | 0.14 | -0.12 | -0.08 | 0.15 | -0.31 | -0.14 |
| % NR<sup>↗</sup> |  |  |  | 0.35 | -0.25 | 0.48 | 0.27 | -0.05 | 0.53 | 0.35 | -0.17 | 0.51 |
| **SHS additions** |  |  |  |  |  |  |  |  |  |  |  |  |
| Irregularity<sup>↓</sup> (duration) |  |  |  |  |  |  | -0.27 | -0.42 | 0.15 |  |  |  |
| Onset latency<sup>↓</sup> |  |  |  |  |  |  | -0.11 | 0.26 | 0.33 | -0.06 | 0.34 | 0.12 |
| Fragmentation<sup>↓</sup> |  |  |  |  |  |  | -0.44 | 0.36 | 0.11 |  |  |  |
| WASO<sup>↓</sup> |  |  |  |  |  |  | -0.32 | -0.01 | -0.28 |  |  |  |

↑ = Higher is better; ↓ = lower is better; ↗ = generally higher is better, but overexpression can be problematic (eg long sleepers, excessive % NR)

## Discussion

There is growing interest in considering sleep health as not merely the absence of a disorder but as a summary of the positive attributes of healthy sleep. Similarly, there is movement towards using a conceptual framework that articulates sleep as multidimensional, potentially summarized as a composite measure (1). We further suggest the value in considering the intrinsic correlations and interactions among sleep measurements that comprise global sleep scores; the challenges in developing optimal composite indices; and the potential value in extending sleep health scores to include additional dimensions that describe common sleep disorders (especially in middle-aged and older populations), quantify sleep architecture, and consider regularity in not only sleep timing but sleep duration.

In evaluating alternative ways to summarize sleep health in an aging multi-ethnic community sample, we found evidence to support the utility of composite sleep health scores based on the Ru SATED framework as well as scores extended to include measures from PSG and actigraphy. In extending the Ru SATED framework with the additional consideration of sleep architecture (particularly % NR), OSA (AHI), and duration regularity, as well as several measures of sleep efficiency/continuity, we observed that the new composite yielded interpretable principal components with more comprehensive scores evincing higher internal reliability than simpler scores. PCA on original and extended Ru SATED frameworks showed that the individual indices of sleep health for each score aggregated in consistent dimensional patterns. The face validity of the extended SHS as a composite was supported by the empirical finding that it summarized broad systematic shifts (most-least favorable categories; 0-3 vs 9-12) across multiple sleep health metrics, including shifts in the initial components of Ru SATED as well as in duration regularity, AHI, and sleep architecture. Moreover, each composite score varied with race/ethnicity, supporting its utility for describing and monitoring sleep health disparities. Adding additional sleep information also suggested important patterns of association. For instance, timing <u>and</u> duration irregularity showed sleep disparities, which might suggest that assessing determinants or barriers to (ir)regular bed <u>and</u> wake times may be beneficial in reducing racial-ethnic sleep disparities rather than a focus on regular bed or wake times alone (25). Although timing and duration regularity are moderately correlated, only about 31% of the variance of one accounts for the other, suggesting these regularity metrics to be patterned yet distinct phenomena. For example, rotating shift-workers may have consistent duration, yet inconsistent timing. Therefore, in middle-age or older cohorts where sleep disorders and circadian disruption are prevalent, there is value in extending global sleep health assessments with data from PSG and actigraphy. Moreover, metrics of OSA, altered sleep architecture, and sleep duration variability predict adverse health outcomes, cognitive decline and mortality, underscoring the potential utility of these data for informing sleep health assessments (26-28).

Our analyses not only supported use of composite indices, but also concurrent consideration of multiple individual dimensions. For example, we found that the individual “drivers” of differences in sleep health composites differed across race/ethnic groups, consistent with prior work in both adults and the elderly (14). For example, compared to Whites, scores in Blacks were influenced by sleepiness and %NR, while scores in Chinese were influenced by AHI. Identifying individual drivers of sleep health may be particularly important for designing targeted interventions. However, post-hoc analysis of the individual components of a summary score requires cautious interpretation, and confidence intervals and p-values for individual dimensions may be less informative than the pattern and trend among point estimates (2). Below we further discuss some of the challenges and opportunities presented by multidimensional sleep health, and analysis of both composite and individual dimensions.

### Statistical correlation among sleep variables and implications for a composite sleep health approach

We demonstrated that many sleep dimensions (both within and across domains) are inter-correlated, reflecting their intrinsic physiological inter-relationships and potential responsiveness to common stressors. Imagine, for instance, that each dichotomized SHS dimension in Table 1 is represented by a light in a circuit. In the *absence* of dependence patterns, sleep health dimensions would function like lights that burn out in parallel circuits, where health in one dimension does not inform the others. However, if there are dependencies, much like a complex circuit with both serial and parallel components, there is increased likelihood of some lights burning out (% NR, sleepiness, sleep fragmentation) if certain other lights burn out (AHI). For instance, in our data, higher sleep irregularity (timing and duration) and less sleep duration tended to co-occur. If this pattern indicates the effects of an underlying common cause or latent factor, including measures of both irregularity and duration should better characterize this driver within the distribution of a composite.

In scales such as the SHS, therefore, the distribution of the composite score may reflect relationships among the individual items that are informative. However, it is challenging to not only select which measures to include in composites, but to determine the optimal way to aggregate and weight each item, and whether to use continuous metrics or cutoff values (discussed later). In choosing sleep variables for a composite, interpretability and ease of data collection argue for a more parsimonious approach; on the other hand, there are potential benefits of more comprehensive inclusion. First suppose that sleep health dimensions have independent errors (e.g. from different measurement modalities, or measurements on independent nights). Analogous to the case that adding test items improves measurement in classical test theory, including additional dimensions within the same domain would reduce measurement error, increasing signal and stability for an underlying latent target. On the other hand, if multiple variables representing the same domain are calculated from the same underlying data, the errors will not be independent; but if it is not known in advance which function/non-linear transformation results in a measure more correlated with other variables in the composite, including both forms may improve the chance of discovering these relationships. Data-driven techniques may allow room for new dimensions/patterns to emerge (29), perhaps better representing underlying drivers, etiological factors, or groups for stratified interventions.

### Public health: Population sleep health assessment by defining optimal ranges and characterizing individual dimensions and patterns across dimensions

For public health utility, however, a categorical basis using prior knowledge to define optimal ranges for each sleep dimension may also be appropriate. Early work operationalized Ru SATED domains by dichotomizing daily diary, questionnaire, or actigraphy measures and summing them into an overall sleep score (21-23). While continuous sleep exposures or outcomes offer greater statistical power, categorical assessments – those involving a cut-point – provide three practical benefits (30). First, cut-points aid clinicians in making decisions. Second, prevalence can be defined and then used for needs assessment: e.g., approximately one third of Americans do not meet sleep quantity recommendations (31). Third, cut-points enable quantification of prevalence trends over time and across groups. Additionally, cut-points may help avoid issues of non-linearities in exposure-response relationships, and cut-points are useful for setting goals for public health initiatives. Categorical assessments may be appropriate even if there is no evidence of a latent, *internal* discrete structure (taxon) for that dimension. As Kessler (2002) notes, “there appears to be no taxon for high blood pressure,” and yet cut-points for blood pressure based on *external* criteria such as risk of stroke guide clinical and public health decisions (30).

Implications of the categorical approach for population sleep health are illustrated in Table 1. At the dimension level, needs assessment becomes clearer. That WASO ≤20 minutes, for instance, only has 6.8% favorability in MESA may suggest a calibration of the NSF’s optimal ranges through extra data collection or may indicate an unmet need in middle-aged and older adults (or both). Both cases are of public health import because age-appropriate optimal sleep ranges can provide a set of public health targets to be validated against external criteria, whereas unmet needs in a community suggest a need for investigation and intervention.

### Leveraging information from both composite scores and individual dimensions: patterns of association illuminating potential sources of sleep health disparities

Our results indicate how composites and individual dimensions can be mutually informative. Compared to Whites, and consistent with prior evidence in MESA which included both adults (aged 54-64 years) and the elderly (65-93 years) (14), adult Blacks in MESA show i) lower sleep health composite scores, on average by more than a full component’s difference for the SHS; and ii) the largest dimension-level disparities for Blacks are in regularity (timing and duration), duration, % N3, and continuity (Figure 2). Viewed from the sleep health paradigm, Figure 3 suggests the likelihood of observing multiple disparities given a large disparity in, and high correlation with, a single sleep dimension. We know why maintenance efficiency (SME) and fragmentation (FI) show a similar pattern of disparity: each measures different facets of sleep continuity. The bases for inter-relationships among sleep irregularity, duration, and continuity and their racial-ethnic patterning are less clear. If timing irregularity is accompanied by circadian misalignment (32), the two-process model of sleep suggests that circadian misalignment and duration variability may interact, suggesting *internal* (to sleep health) causal relationships. On the other hand, the observed disparities may reflect *external*, socioecological causal factors such as light, shiftwork, etc. that influence sleep regularity, duration, and continuity. Here, inter-correlation is cast as a source of research hypotheses about patterns of multiple dimensions of sleep (2).

Consequently, we offer an interpretation of Buysse’s definition that sleep health is a *pattern* of sleep-wakefulness, illustrated by the dimensions of the SHS. Sleep health researchers could estimate independent models for each dimension and assemble the interpretation of these models in light of i) a global sleep health effect; and ii) correlated sleep dimensions. For instance, individuals of Hispanic ethnicity showed average global sleep health (as indexed by the SHS) that was significantly lower than that of Whites (Figure 2a). However, few of the dimension-level disparities for Hispanics are distinct from the reference. The old paradigm that focused on single dimensions may have under-estimated Hispanic sleep disparities. Rather, under the sleep health paradigm, it is notable that the Hispanic sleep health disparity *pattern* mostly resembles that of Blacks.

### Statistical correlation among sleep variables: patterns of association and implications for intervention targeting and evaluation

Changing a single sleep dimension may have “carry over” effects on other sleep attributes. For example, an intervention which improves sleep regularity may improve sleep continuity. Conversely, if sleep deteriorates in one dimension (e.g. duration), deteriorations in other dimensions of sleep health may be expected (e.g. sleepiness, satisfaction, duration variability, timing variability, etc.). For dimensions more difficult to directly target (% NR), interventions could focus on correlated dimensions that are amenable to intervention (e.g., irregularity, AHI). Examining multiple dimensions also may be useful in evaluating the effects of interventions such as zolpidem, which may positively influence features such as sleep latency and WASO but negatively influence sleepiness/alertness and NR sleep. Interventions that affect only one sleep dimension may be the exception rather than the rule.

Multidimensionality opens the possibility of ‘indirect’ or ‘root-cause’ targeting, such as targeting OSA to improve multiple sleep health metrics such as alertness, quality, continuity, and % NR on a population level. Indeed, the high prevalence of OSA, its underdiagnosis, and causal effects on other sleep dimensions suggest OSA as a driver of poor sleep health (19, 33-35), and thus an attractive intervention target to improve multiple sleep health dimensions (36-38). Notably, multidimensionality does not preclude focus on multiple dimensions. (39, 40)

While sleep duration has been a key focus of epidemiological research, our analyses support the importance in examining the contributions of multiple dimensions. Findings on the detrimental effects of insufficient sleep duration may implicitly capture effects of poor sleep in correlated dimensions – such as regularity, continuity, sleep architecture, OSA, placement in the 24-hour day, and sleepiness/alertness – and erroneously attribute this omnibus effect to duration. Investigating dimensions other than duration may thus inform interventions on a population scale. It may prove more accurate to say: ‘Sufficient sleep duration (*and all that it implies*) is protective of later health.’ Alternatively, ‘Sleep health, of which average sleep duration is one vital component, is protective of later health.’ Novel sleep health analyses using machine learning support this conclusion (7, 41).

## Limitations

The first limitation concerns the dependencies among dimensions, which may be one of the more interesting and useful features of sleep health. The source(s) of these dependencies, however, remain poorly understood. It is possible that the correlations observed are particular to the sample or attributable to confounding factors such as the socioecological context, age or other factors.

A second limitation concerns evolving or uncertain definitions and criteria of sleep health. Consensus measures to assess sleep health with objective sleep data are lacking. We leveraged comprehensive sleep data in MESA, and in doing so traded the parsimony of the original Ru SATED framework for flexibility in adding dimensions empirically linked to race-ethnicity (42, 43) as well as downstream health (26, 44) – at the cost of scalability. Future work might estimate weights for each dimension in relation to a health outcome, with the limitation that weights may be particular to the population and outcome.

The composites used self-reported and objective sleep indices. Even for the simple Ru SATED score, which we derived by using objective data to define timing, duration and efficiency, and from questionnaire data on quality and sleepiness/alertness, the PCA showed that the objective and subjective measures each loaded on distinct components. These distinct loading could reflect measurement bias rather than dimensional differences. The correspondence between purely self-reported composite scales and those involving objective measures is not characterized, and prior research indicates systematic differences in estimates between self-report and objective measures according to short/long duration, sleep efficiency, health, and socio-demographics (45).

Moreover, explicit guidance on population specific thresholds is limited. The core definition of Ru SATED dimensions is evolving as well, with sleep regularity a recent addition. A canon of sleep health parameters for evolving measures of sleep micro-architecture (e.g. spindles, k-complexes) has yet to be established despite emerging data on the unique information contained in these measures (27, 46).

These limitations highlight that sleep health is undergoing conceptual advances and scale development (1, 6), scale validation (5, 6), implementations of sleep health in cohort studies and community samples (21, 47), innovations in methodological approaches (4, 7, 41, 48), optimal range and threshold determination (18, 22), and many practical choices on how best to analyze multidimensional sleep data. We suggest that statistical dependencies are an additional factor to contend with. As sleep health continues to develop as a field, we suggest two solutions that may help sleep health’s future development.

### Solution 1: Studying challenges and solutions in disciplines which have already shifted to multidimensional paradigms

From multidimensional/symptom psychiatric epidemiology (30), it is instructive to observe the outcome of the tension between epidemiological theories of ‘distribution shifts vs targeted intervention’ and policy making (30). Although shifting distributions is a powerful public health approach, the current policy paradigm favors targeted intervention, prevalence estimates, reporting prevalence trends over time, etc. (31). Thus, identifying an initial threshold target, with monitoring of impact of interventions on this target and related health outcomes, provides an example of how sleep health research can be translated to policy. This approach is similar to the long-standing practices of using blood pressure cut-offs as public health and individual targets despite evolving data on specific thresholds that confer disease (30). Future research to develop normative sleep ranges will further support the utility of this approach for policy makers and the public health. Logically implied are similar needs for developing and validating normative or optimal ranges for composite sleep metrics.

Advantages to using composite scores include the reduction of multiple testing burden (as compared to analysis of multiple individual dimensions) and the ability to detect the effects of multiple small effects. An analogy can be drawn between sleep health and nutrition, in which most nutrients appear to contribute in small ways to a larger composite effect (and sometimes this composite effect better describes nutritional profiles than when individual nutrients are examined) (2). Given their simplicity, composites also have ready public health application, similar to promoting diets such as the Mediterranean diet rather than recommending specific nutrient targets. (2). However, while summary scales may be selected *a priori* based on extant empirical or conceptual work, prospective data are needed to demonstrate that they predict clinically meaningful outcomes.

Methodological advances in sleep health composites are anticipated with advances in sleep knowledge (and vice versa), perhaps leveraging increasingly larger datasets (49). Nonetheless, there is a need to recognize that the components of global sleep health scores will likely vary according to the questions at hand (e.g., interest in a pediatric versus aging population; interest in cardiovascular disease or cancer). Future work might draw from prior literature on composites in other fields which also deal with complex systems with high-dimensional data. This may be of high utility in choosing how to weight and aggregate the complex set of phenomena comprising sleep health in a context-specific way (50).

Another tension appears to depend on sleep health’s complexity and wide-ranging importance. Because sleep health is multidimensional, whose dimensions may be variably more/less responsive to a wide set of individual and social factors, and whose different components may be more/less consequential for different outcomes, a single, canonical operationalization of sleep health may be suboptimal for any particular project. It may be useful to construct at least two sleep health indices: i) canonical Ru SATED (one dimension per domain); and ii) modified Ru SATED for a specific project (if necessary). This may help preserve consistency in the field while also affording flexibility in answering varied research questions. For assessing racial-ethnic disparities, the Sleep Health Score showed adequacy in distinguishing between groups; analysis of individual effects pointed to the strongest and modifiable drivers of the composite association, for public health targeting (regularity, duration).

### Solution 2: Objective and longitudinal sleep data

A recent NIH workshop report on sleep health disparities emphasized that multidimensional sleep health must be experienced on a *regular* basis (51). However, there are little extant cohort data to assess whether sleep health is experienced on a regular basis, and what factors produce regular sleep health. Moreover, there are few data on within-individual correlations of sleep dimensions across time, and thus little understanding of the ‘natural histories’ of good or poor multidimensional sleep health, and a lack of longitudinal data limits causal modeling through natural experiments. Two solutions to these problems are 1) analysis of multi-day sleep studies as a time series (52); 2) longitudinal, comprehensive cohort sleep data collection.

First, Hale, Troxel, and Buysse (2020) argued that daily sleep assessments, modeled as a time series, may unpack bi-directional relationships between sleep and regular exposures such as diet and stress (52). Such work can help to capture the effect of short-term exposures. Moreover, a single night’s poor sleep health in a particular dimension may cascade to affect other dimensions the same or next night or may suggest common exposures that affect multiple dimensions (e.g. rotating shift work) (9). Multi-day comprehensive sleep assessment with daily physical and psychosocial exposure measurements should elucidate how sensitive sleep health dimensions may be to acute, fluctuating exposures. Thus, future cohort studies might consider daily psychosocial instruments or an expanded sleep diary(now possible through electronic data collection) to capture daily exposures of interest across the sleep study period and consider how such data are summarized as composites or individual dimensions.

Consumer wearables are a promising alternative to traditional actigraphs. Consumer wearables are relatively inexpensive and are amenable to data collection over long periods. Sleep data can be paired with physical activity measurement and geo-coding to get a fuller picture of sleep-wake patterns.(53) As these data are collected on larger numbers of individuals, it will be important to consider whether they provide novel metrics of sleep timing, variability and others features not apparent in shorter-term measurements.

Longitudinal sleep data are critically needed to better understand the ‘natural history’ of multidimensional sleep health, and may help identify which dimensions are the most variable over long time scales and how favorable patterns of sleep health may become distorted by changes in behavior or environment over the life course (e.g. major life events) (54, 55). Accordingly, longitudinal data may help better characterize critical features and etiology of consequential sleep health phenotypes. For instance, Vgontzas et al. (2012) reported that objective short sleep duration (<6 hours) at baseline was associated with higher odds of persistent insomnia 7.5 years later compared to normal sleepers and those with fully remitted insomnia (56). Longitudinally collected sleep phenotypes can clarify time-dependent features whose proximal cause and age of onset and time-dependent modifying factors are otherwise difficult to characterize.

Longitudinal data may be particularly useful to clarify specific sleep drivers and levers. We suggest sleep regularity (in timing and duration) as candidates for further longitudinal research due to their i) potentially modifiable nature (as reflected in sleep hygiene recommendations for consistent bed and wake times), ii) consistent correlation with many other sleep dimensions (Figure 3), and iii) ability to forecast incident metabolic dysfunction and cardiovascular events (12, 13). If regularity as a candidate driver of sleep health is supported by longitudinal studies, then targeted interventions can be tested, including the impact on sleep health composites and clinical endpoints.

In summary, comprehensive and repeated sleep assessment may inform the etiology and ‘natural history’ of multidimensional sleep health, identify candidate drivers and phenotypes, and help establish normative sleep health ranges for scientific knowledge and as targets for public health. To create robust sleep health scores, additional research showing their value in predicting health outcomes is needed.

### Incorporate qualitative data

Finally, qualitative research complements other research methods, providing information, from the participant’s point of view, about: i) barriers and facilitators to achieving sleep health and ii) impact of sleep on overall health and well-being. For instance, in a small Boston sample of racially-ethnically diverse low-income adults, commonly stated barriers to “good sleep” were work (or multiple) work schedules, consuming large quantities of soda in the evening, reluctance to stop using personal electronics at bed-time, child-care, and financial worries (25). Further investigation of factors such as these can be thoughtfully integrated into future data collection efforts that inform multidimensional sleep. Qualitative work is vital to retrospectively understand changes in sleep health and is thus of high value to understanding etiology. For instance, one of the richest pieces of evidence in support of the ‘precipitation’ component of the 3-P model of insomnia (predisposition, precipitation, perpetuation) comes from Healey et al. (1981) who used a set of retrospective interviews assessing stressful life events as precipitants of insomnia complaints (55).

### Summary

Multidimensional sleep health represents a paradigm shift in sleep science. Two useful frameshifts are that sleep health is both conceptually and operationally a composite concept and that sleep dimensions do not exist in isolation. These frameshifts logically implicate sleep disordered breathing and sleep regularity as targets for further research and intervention. To fully explore the implications of this new paradigm, comprehensive longitudinal assessment of sleep is necessary.

## Data Availability

The secondary, de-identified data used in the manuscript are available at the National Sleep Research Resource (NSRR): https://sleepdata.org/

## Acknowledgements

This research was supported by National Institute of Health grants 5T32HL007901-14, R35HL135818, R01HL098433; and by contracts HHSN268201500003I, N01-HC-95159, N01-HC-95160, N01-HC-95161, N01-HC-95162, N01-HC-95163, N01-HC-95164, N01-HC-95165, N01-HC-95166, N01-HC-95167, N01-HC-95168 and N01-HC-95169 from the National Heart, Lung, and Blood Institute.

## Declaration of conflicts of interest

JC, MW, TH, SB, and SR report funding from the NIH. SB reports consultant fees from Eisai, Inc, and Merck Sharpe & Dohme. The authors declare no other conflict.

## Human Subjects Protection

This research used secondary, de-identified, publicly available data (National Sleep Research Resource). Participants provided informed consent for the original study, which complied with the Declaration of Helsinki.

## Appendix A. Comparison of Ru SATED and the Sleep Health Score (SHS)

**Table A1.** Compare/contrast Ru SATED and Sleep Health Scores.

| Sleep dimension | Ru SATED | SHS |
| --- | --- | --- |
| Regularity | x | x |
| Satisfaction | x | x |
| Alertness | x | x |
| Timing | x | x |
| Efficiency | x | x |
| Duration | x | x |
| Sleep architecture |  | x |
| Duration regularity |  | x |
| Onset latency |  | x |
| Dimensions | 1 measure per domain<br>(parsimony) | Multiple related<br>measures allowed (see<br>discussion of<br>composite scores) |

**Figure A1.**
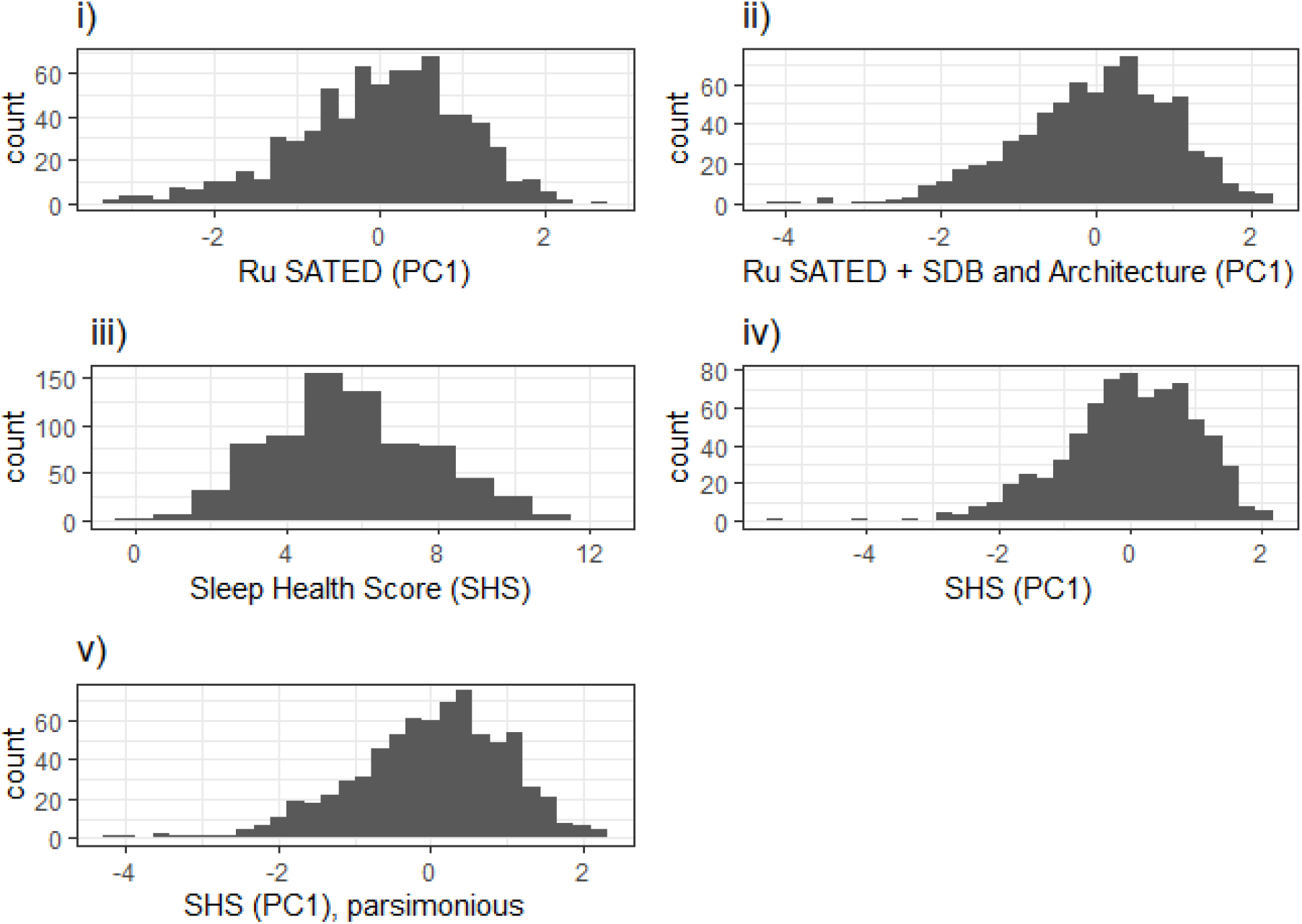
Distributions of sleep health composites: i) Ru SATED (PC1), ii) Ru SATED + SDB and Architecture (PC1), iii) Sleep Health Score, iv) Sleep Health Score (PC1), v) Sleep Health Score (PC1) parsimonious (one measure per domain)

## Appendix B.

**Table B1.** Age and sex-adjusted regression results for Ru SATED (PC1), Ru SATED + SDB/Architecture (PC1), SHS (PC1), and the SHS.

|  | Ru SATeD (PC1) |  | Ru SATeD +<br>OSA/Architecture (PC1) |  | SHS (PC1) |  | SHS |  |
| --- | --- | --- | --- | --- | --- | --- | --- | --- |
| | $\beta$ [95% CI] | p | $\beta$ [95% CI] | p | $\beta$ [95% CI] | p | $\beta$ [95% CI] | p |
| Chinese | -0.28<br>[-0.50, -0.06] | 0.013 | -0.24<br>[-0.46, -0.02] | 0.029 | -0.25<br>[-0.46, -0.03] | 0.027 | -0.29<br>[-0.77, 0.19] | 0.235 |
| Black | -0.87<br>[-1.05, -0.70] | <0.001 | -0.82<br>[-0.99, -0.65] | <0.001 | -0.76<br>[-0.92, -0.59] | <0.001 | -1.34<br>[-1.72, -0.97] | <0.001 |
| Hispanic | -0.32<br>[-0.50, -0.14] | <0.001 | -0.31<br>[-0.48, -0.13] | 0.001 | -0.33<br>[-0.51, -0.16] | <0.001 | -0.77<br>[-1.16, -0.38] | <0.001 |
| Female | 0.29<br>[0.15, 0.43] | <0.001 | 0.54<br>[0.41, 0.67] | <0.001 | 0.53<br>[0.40, 0.67] | <0.001 | 0.55<br>[0.25, 0.84] | <0.001 |
| Age (years) | 0.02<br>[-0.00, 0.04] | 0.077 | 0.00<br>[-0.02, 0.02] | 0.945 | -0.02<br>[-0.04, 0.01] | 0.161 | -0.01<br>[-0.06, 0.04] | 0.796 |
| (Intercept) | -1.04<br>[-2.41, 0.33] | 0.138 | -0.02<br>[-1.36, 1.32] | 0.976 | 0.98<br>[-0.37, 2.34] | 0.155 | 6.36<br>[3.36, 9.36] | <0.001 |

**Table B2.** Variable-level regression results for the components of the SHS. N=735.

| Variable: | Black<br>RR [95% CI] | Hispanic<br>RR [95% CI] | Chinese<br>RR [95% CI] | Age<br>RR [95% CI] | Female<br>RR [95% CI] | (Intercept)<br>RR [95% CI] |
| --- | --- | --- | --- | --- | --- | --- |
| MPsd | 0.44 [0.31, 0.62] | 0.72 [0.54, 0.97] | 0.80 [0.56, 1.14] | 1.02 [0.98, 1.06] | 0.97 [0.77, 1.22] | 0.12 [0.01, 1.19] |
| TSTsd | 0.42 [0.33, 0.55] | 0.85 [0.71, 1.02] | 0.79 [0.62, 1.01] | 1.00 [0.97, 1.02] | 0.92 [0.79, 1.07] | 0.61 [0.13, 2.96] |
| Fragmentation | 0.55 [0.42, 0.73] | 0.68 [0.52, 0.88] | 0.90 [0.68, 1.19] | 0.97 [0.93, 1.00] | 1.57 [1.27, 1.95] | 2.09 [0.29, 15.32] |
| TST | 0.65 [0.57, 0.74] | 0.80 [0.72, 0.90] | 0.76 [0.65, 0.88] | 0.99 [0.98, 1.01] | 1.12 [1.02, 1.23] | 0.99 [0.39, 2.52] |
| % N3 | 0.56 [0.35, 0.88] | 0.62 [0.39, 1.00] | 1.09 [0.68, 1.75] | 0.97 [0.92, 1.03] | 1.80 [1.25, 2.59] | 0.45 [0.02, 12.82] |
| SME | 0.83 [0.76, 0.92] | 0.89 [0.81, 0.98] | 0.85 [0.74, 0.96] | 1.00 [0.99, 1.02] | 1.17 [1.08, 1.27] | 0.60 [0.28, 1.28] |
| AHI | 0.99 [0.86, 1.15] | 0.88 [0.75, 1.03] | 0.83 [0.67, 1.03] | 0.97 [0.95, 0.99] | 1.71 [1.48, 1.97] | 2.44 [0.74, 7.99] |
| SOL | 0.91 [0.81, 1.01] | 0.85 [0.75, 0.96] | 1.08 [0.95, 1.21] | 1.02 [1.00, 1.03] | 0.83 [0.76, 0.91] | 0.29 [0.12, 0.73] |
| % NR | 0.75 [0.60, 0.95] | 1.09 [0.89, 1.33] | 1.01 [0.77, 1.31] | 1.00 [0.97, 1.03] | 1.29 [1.08, 1.54] | 0.34 [0.07, 1.82] |
| ESS | 0.89 [0.83, 0.96] | 0.99 [0.93, 1.05] | 1.01 [0.93, 1.09] | 1.01 [1.00, 1.02] | 1.01 [0.96, 1.06] | 0.45 [0.26, 0.78] |
| Timing | 0.90 [0.79, 1.01] | 0.92 [0.81, 1.04] | 1.14 [1.00, 1.29] | 0.99 [0.97, 1.00] | 1.02 [0.93, 1.12] | 1.31 [0.51, 3.35] |
| WASO | 0.63 [0.33, 1.20] | 0.98 [0.56, 1.72] | 1.37 [0.74, 2.56] | 0.90 [0.83, 0.97] | 1.71 [1.05, 2.77] | 31.38 [0.34, 2909.80] |
| Quality | 1.10 [0.90, 1.35] | 0.93 [0.74, 1.17] | 1.30 [1.03, 1.64] | 1.05 [1.02, 1.08] | 0.91 [0.77, 1.06] | 0.02 [0.00, 0.12] |

## Notes

### Author Declarations

Primary data collection was approved by the Institutional Review Boards of MESA field centers. Publicly available, de-identified data were used in secondary analyses. Data are available at the National Sleep Research Resource (NSRR): https://sleepdata.org/

